# Metabolic and immune markers for precise monitoring of COVID-19 severity and treatment

**DOI:** 10.1101/2021.09.05.21263141

**Authors:** André F. Rendeiro, Charles Kyriakos Vorkas, Jan Krumsiek, Harjot Singh, Shashi Kapatia, Luca Vincenzo Cappelli, Maria Teresa Cacciapuoti, Giorgio Inghirami, Olivier Elemento, Mirella Salvatore

## Abstract

Deep understanding of the SARS-CoV-2 effects on host molecular pathways is paramount for the discovery of early biomarkers of outcome of coronavirus disease 2019 (COVID-19) and the identification of novel therapeutic targets. In that light, we generated metabolomic data from COVID-19 patient blood using high-throughput targeted nuclear magnetic resonance (NMR) spectroscopy and high-dimensional flow cytometry. We find considerable changes in serum metabolome composition of COVID-19 patients associated with disease severity, and response to tocilizumab treatment. We built a clinically annotated, biologically-interpretable space for precise time-resolved disease monitoring and characterize the temporal dynamics of metabolomic change along the clinical course of COVID-19 patients and in response to therapy. Finally, we leverage joint immuno-metabolic measurements to provide a novel approach for patient stratification and early prediction of severe disease. Our results show that high-dimensional metabolomic and joint immune-metabolic readouts provide rich information content for elucidation of the host’s response to infection and empower discovery of novel metabolic-driven therapies, as well as precise and efficient clinical action.

## Introduction

The pandemic caused by infection with the severe acute respiratory coronavirus type 2 (SARS-CoV-2) has infected more than 218 million people worldwide as of August 2021, caused more than 4.5 million deaths^1^, and strains health systems on an unprecedented scale. The most common manifestations of COVID-19 are fever, cough, and dyspnea^2,3^, but thromboembolic events and other organ involvement are also common in patients with severe disease^2,4,5^. Molecularly, severe COVID-19 disease is characterized by uncontrolled inflammatory syndrome caused by immune system hyperactivation^6–11^. The most effective treatments are thus based on general immunosuppression with glucocorticoids^12^ or neutralization of the pro-inflammatory interleukin 6 (IL-6) with tocilizumab^13^.

Several laboratory tests such as albumin^14^, CRP^15^, lymphocyte abundance^16–19^, IL-6^20^, and the fibrin degradation product D-dimer^21^ have been used to monitor COVID-19, with their levels variably associated with disease severity. While these routinely available assays may have some clinical use in disease prognostication, they depict an incomplete landscape of pathophysiological changes associated with COVID-19. However these tests are mostly a readout of the inflammatory state and do not capture a wide but still interpretable view of the physiological state of COVID-19 patients. One possible approach is to increase the dimensionality of the system by the use of mixed-modality profiling such as the combination of immune population quantification and circulating cytokine levels^22,23^. While much work has been done on the characterization of the host immune response through cytometric or serological methods, characterization of the metabolic state of COVID-19 patients has just begun^24–26^.

There are several lines of evidence demonstrating the importance of metabolic species - in particular lipids - during viral infection. Lipids are structural components of the host’s cellular and organellar membranes, taking an active role in crucial cellular functions such as molecular trafficking, but are also of importance during viral attachment, internalization, packaging and release^27,28^. In animal models, it has been shown that cholesterol composition of membrane lipid rafts underpins the infectivity of SARS-CoV-2^29^. Beyond its structural functions, lipids are also crucial for energy supply and intracellular signalling^30,31^. It is plausible that viral-induced changes in host metabolism during COVID-19 in metabolites such as glucose and lipids^32^ may be beneficial for the infection, by altering intracellular signaling and conditioning immune response. The host metabolome - lipids in particular - have therefore been proposed as potential biomarkers of COVID-19 disease severity^33^, and as therapeutic targets to counteract excessive immune activation. Nuclear magnetic resonance (NMR) spectroscopy applied to the measurement of metabolites provides a great balance between precise and reproducible measurements, the breadth of analytes measured, and the logistical efforts necessary for data production^34–36^. Importantly, it is also capable of discerning different lipid species in circulating lipoprotein particles.

In this work, we use NMR spectroscopy to identify changes in serum metabolome composition of COVID-19 patients that are associated with disease severity and tocilizumab treatment, and provide a method for precise disease monitoring, patient stratification, and early prediction of severe disease based on joint immuno-metabolic measurements.

## Results

### Longitudinal NMR metabolomics of COVID-19 patient plasma

We conducted an observational study of 75 individuals with acute or convalescent COVID-19 that were treated at New York Presbyterian Hospital and Lower Manhattan Hospitals, Weill Cornell Medicine as in- or out-patients between April and July 2020. The disease was categorized using World Health Organization disease severity scale for the prognostication of COVID-19 patients^37^ (henceforth referred to as “WHO score”), which use clinical events such as patient admittance, amount of supplemental oxygen needed, or the need for mechanical ventilation (**Figure 1A**). Serum samples were collected at hospital admission, when permissible approximately every 7 days thereafter, and for convalescent patients as outpatients at least 90 days from symptom onset (109 samples from 75 patients, 32 convalescent). Of all patients, 35 (47%) presented with low to mild disease severity, and 30 (40%) with moderate to severe disease. We also collected serum from healthy, COVID-19 negative donors (n = 9). The median age of COVID-19 patients was 53 years, which was comparable with that of healthy donors (51 years) (**Table S1**).

**Figure 1:**
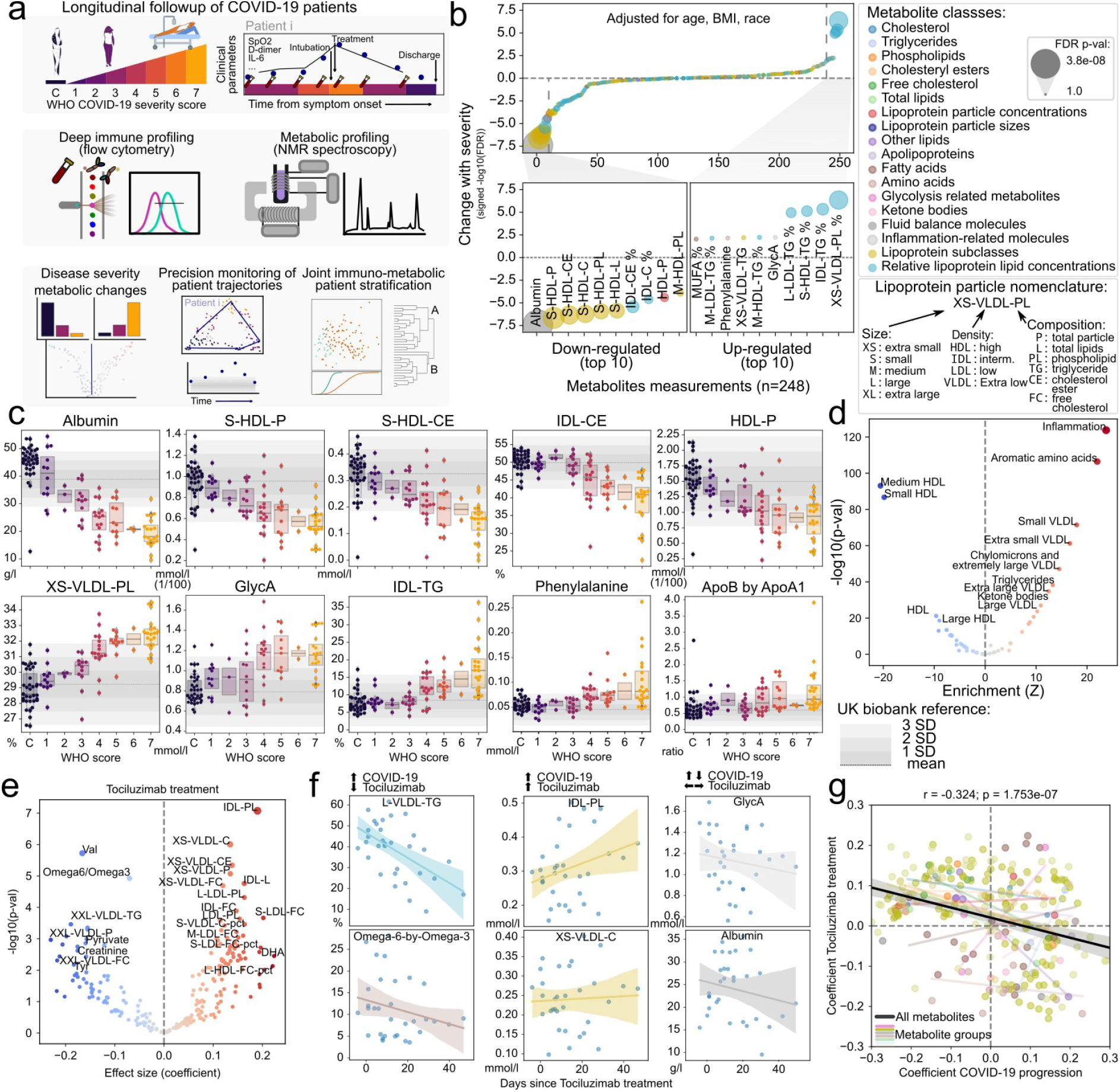
Discovery of metabolic biomarkers of COVID-19 severity and treatment. **a**) Schematic description of the patients under study, data types collected and approaches for their analysis. **b**) Association of metabolite abundance with COVID-19 severity for all 248 metabolic species (upper panel). The lower panel illustrates the 10 metabolites most associated with disease severity for each direction. **c**) Distribution of metabolite abundance for the metabolites most associated with COVID-19 severity depending on the sample WHO score classification. The grey horizontal dashed line represents the mean abundance of the metabolite in over 150,000 individuals from the UK biobank cohort and grey bars represent the standard deviation from the mean. **d**) Enrichment analysis of metabolites changing with COVID-19 severity in functional terms. **e**) Association of metabolite abundance with the time since tocilizumab treatment. The coefficient values refer to the change per day in relation to the mean. **f**) Abundance of metabolites with discordant (left), concordant (center) or indifferent (right) change between COVID-19 severity and tocilizumab treatment for treated patients. **g**) Comparison of the coefficients of change in COVID-19 severity (x-axis) and effect of tocilizumab treatment over time (y-axis). Each point is a metabolite colored by its class identity as in b). The black regression line indicates the overall trend between all metabolites, while the colored regression lines indicate the trend for each group of metabolites as in panel b).

We performed targeted high throughput NMR-based detection of metabolites in circulating blood serum (Nightingale Health Ltd.) (**Figure 1A, Table S2-3**). The NMR assay detected 168 metabolite species in absolute molar quantities, and 81 additional measurements of relative proportions covering diverse metabolic species such as lipids and fatty acids, apolipoproteins, amino acids, ketone bodies, and other molecules with known prognostic value across various diseases such as albumin, creatinine, and apolipoprotein levels^38–40^ (**Figure S1a-c**). The panel is dominated by the diversity of lipids and by lipoprotein-associated lipid species which were fractionated based on their relative density and size (**Figure S1d-f**). Overall, measurements of the metabolic species had excellent reproducibility and high signal-to-noise ratio (**Figure S1g-h**). Upon relating the abundance levels of all metabolite species across all samples, we find that metabolites were heavily co-regulated (**Figure S1i**).

### Metabolic changes associated with COVID-19 severity and treatment

In order to identify the metabolic features associated with COVID-19 outcome, we leveraged linear mixed effect models to explain COVID-19 disease severity as a function of metabolite levels independently from patient age, gender, race, and body mass index (BMI) (**Figure 1b, Figure S2a-b**). While most of the 249 metabolite species known prognostic value across various diseases such as albumin, creatinine, and apolipoproteins showed no association with disease severity as measured by the WHO score, we found significant associations for 56 metabolites (p < 0.05, adjusted for multiple testing with the Benjamini Hochberg False Discovery Rate (FDR) method), which were dominated by lipid and lipoprotein subclasses (**Table S4**). Specifically, we found that Albumin, high-density lipoprotein (HDL) and small HDL particle species, as well as the cholesteryl-ester component of HDL and intermediate-density lipoproteins (IDL) declined proportionally with the increase in WHO score, with steeper decline in the most severe cases (**Figure 1c**). On the other hand, extra small, very low-density lipoprotein (VLDL) particles with increased phospholipids component and extra-small VLDL, IDL, LDL and HDL with increased triglycerides were correlated with increased severity. Additional variables associated with increased disease severity were the acetylated glycoproteins (GlycA) - a spectroscopic marker of systemic inflammation^41^, phenylalanine, and fraction of monounsaturated fatty acids (MUFA). We also observed a significant association of acetoacetate, and the ratio of apolipoprotein B to A1 (ApoB/ApoA1) to disease severity (**Figure S2c**). Of note, the levels of many of the mentioned metabolites in severe COVID-19 (WHO score 4-7) were higher by more than three standard deviations than the mean of a large non-COVID population from the UK Biobank (150,000 samples)^42,43^, illustrating the degree of metabolic disarray in the serum of COVID-19 patients with severe disease. These results are also in agreement with previous reports^24,44^.

Additionally, we performed enrichment of the changes associated with COVID-19 severity in metabolite groups based on their biophysical properties and known physiological roles. This analysis revealed increased levels of inflammation markers, amino acids and triglycerides, but above all confirmed the deep unbalance in lipoprotein composition, size, and density (**Figure 1d, S3a**), where increased severity is associated with decreased lipoprotein density and increased size, which are in line with the increased triglyceride content of the particles. One exception is extra-small VLDL particles (3-6 nm) which are also increased in severe disease. Taken together, the observed changes reveal considerable metabolic changes in COVID-19 patients dependent on disease severity. As a comparison, we investigated the association of routinely collected clinical biomarkers with COVID-19 severity in our cohort and found that only lactate hydrogenase (LDH) was significantly associated with disease severity (**Figure S3b-c**), while biomarkers of overall metabolic homeostasis such as aspartate aminotransferase (AST) and alanine aminotransferase (ALT) were not.

Among the therapeutic options for COVID-19, Tocilizumab, an inhibitor of the pro-inflammatory interleukin-6 (IL-6) was used in COVID-19 patients with elevated inflammatory markers and rapidly escalating oxygen requirements. In our cohort, 10 (12%) patients were treated with Tocilizumab. To assess metabolic changes associated with tocilizumab treatment, we fit a linear model on the time since treatment with age, gender, race, BMI, and disease severity as covariates. Several metabolite species were significantly associated with tocilizumab treatment (**Figure 1e** and **Figure S4a-c, Table S5**), in particular an increase in VLDL particles but also in their cholesterol content (both free and esterified), reduction of valine levels, triglyceride content of VLDL, and ratio of the polyunsaturated fatty acids (PUFA) Omega 6 to 3 - a ratio associated with the pathogenesis of various diseases^45,46^ (**Figure 1f**). However, we also observed metabolite species that were significantly changed with COVID-19 severity with no apparent change with tocilizumab treatment (**Figure 1e-f**, and **Figure S4e-g**). Across all metabolite species, we observed a trend for patients treated with Tociluzumab to have a metabolic state more similar to patients with milder disease over time (**Figure 1g**), which suggests that the administration of Tociluzimab could contribute to a partial rescue of some of the effect of severe disease on the metabolism of COVID-19 patients.

### Precise monitoring of COVID-19 clinical trajectories by intra-patient metabolome dynamics

Given the sensitivity of targeted NMR metabolomics to detect changes of disease severity in COVID-19 patients, we hypothesized that these data could be used as a rich, multivariate measurement of disease severity grounded in metabolic data. First, to understand the temporal dynamics of the metabolism of COVID-19 patients, we created a two-dimensional latent space using the abundance of the metabolites across all samples (**Figure 2a**). This space was largely driven by the severity of disease and clinical outcomes associated with it such as hospitalization, intubation and death, and the time since symptom onset (**Figure 2a** and **Figure S5a-b**). This allowed us to use the distribution of clinical attributes on the space to inform of the relative risk of adverse outcomes (including hospitalization, intubation and death) for patients (**Figure 2a, right** and **FigureS5c**). Importantly, this relationship is largely independent of the statistical method used to construct a latent space (**Figure S5d**). Second, we used pseudotime to derive a direction of progression through the latent space for each patient over multiple timepoints. This allowed us to order samples based on their predicted trajectory along the overall course of disease severity (**Figure 2b**). Finally, we were able to observe the trajectory of each patient along the latent space, and by weighting the amount of change by the time between timepoints we could derive a measure of speed of change of metabolome for each patient, and an aggregate measure of how much the metabolome changed over time (**Figure 2c-e**). For example, patients 23 and 24 have similar trajectories at start - starting at an area of intermediate risk and progressing to an area of highest severity -, but later move to an area occupied most by healthy and convalescent individuals or remain in the area of highest risk, respectively (**Figure 2d**). These divergent trajectories are apparent in the predicted relative risk from metabolic data, while a single marker such a GlycA tends to inform only of one specific aspect of the metabolome (inflammation) (**Figure 2e**).

**Figure 2:**
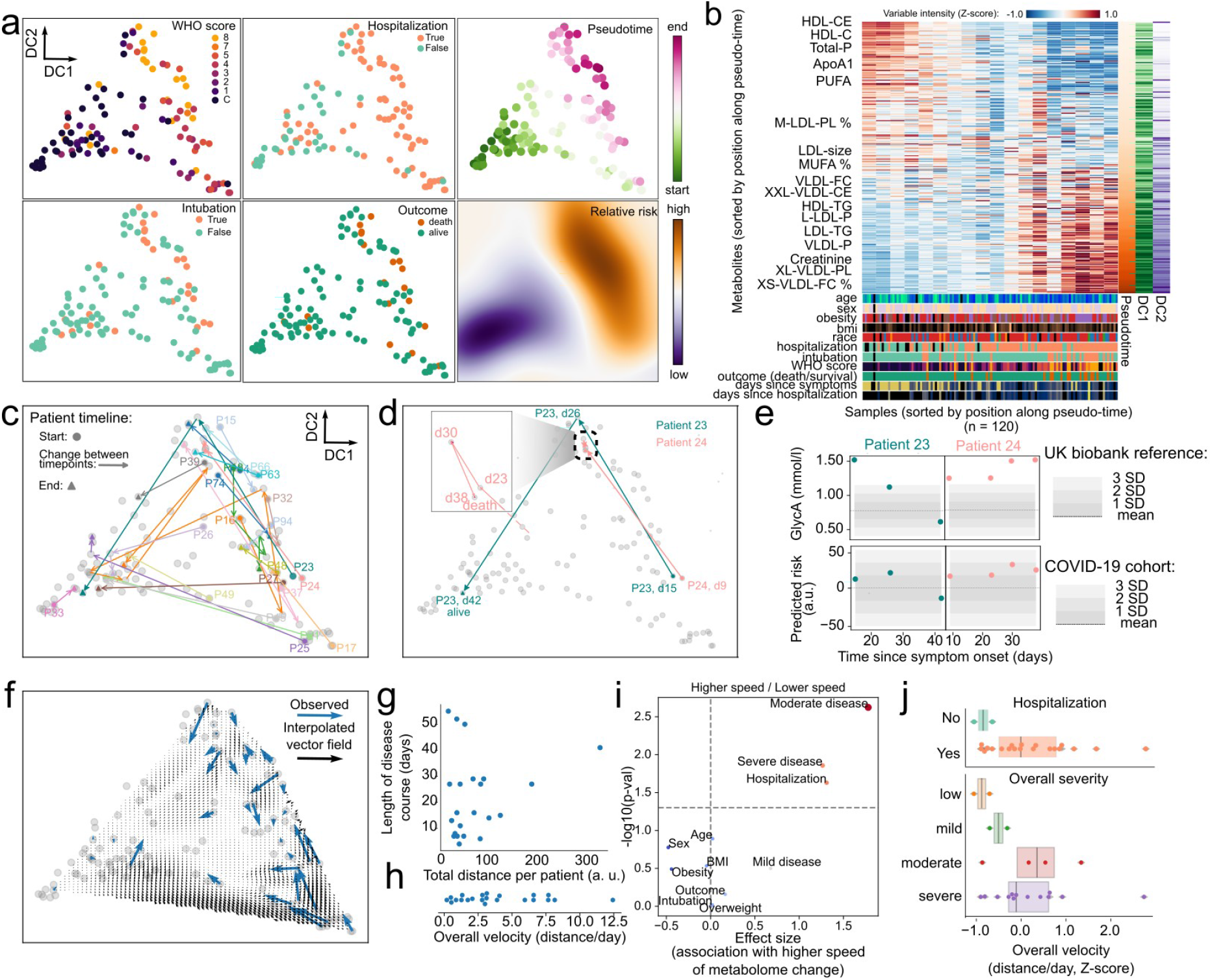
Use of high-dimensional metabolic data for precise disease monitoring. **a**) Latent representation of metabolic data for all samples in two dimensions using diffusion components. In the first two panel columns samples are colored by their value of WHO score, whether the patient was hospitalized, intubated and their survival. The rightmost column indicates the position of each sample within the inferred pseudotime and the relative risk for the whole two-dimensional space. **b**) Heatmap with relative abundance of metabolites for all the samples where both axes are sorted by their relative position along the inferred pseudotime. The lower part of the plot indicates the values of clinical parameters for every sample. **c**) Trajectory of each patient across the latent space during their clinical course. Patients with at least three samples are colored distinctly while the remaining are colored in gray. **d**) Particular trajectories for patients 23 and 24 as in c). The inset illustrates the stagnated course of patient 24. dN = n days since symptoms onset. **e**) Values of GlycA and the predicted risk for patients 23 and 24 along the clinical trajectories of each patient. The shaded area in the GlycA plots represents the distribution of that metabolite in the UK biobank cohort, while the shaded area for predicted risk represents the distribution of the COVID-19 cohort. **f**) Vector field of velocities in the latent space interpolated from the observed velocity vectors for all patients (blue). **g**) Relationship between total distance moved per patient in the latent space over the whole clinical course and its length in days from symptom onset. **h**) Distribution of average velocities across the whole clinical timeline for every patient. **i**) Association analysis between clinical variables and the average velocity of each patient. *p*-values have been adjusted with the Benjamini-Hochberg FDR method. **j**) Illustration of differences between patient velocities and their hospitalization or overall disease severity across the whole clinical timeline.

The longitudinal aspect of the data and its reduction to a single landscape further allowed us to study the temporal kinetics of disease severity and recovery (**Figure 2f**). We hypothesized that the overall speed of each patient along their timeline could be related to their clinical status. We observed that different patients can have largely different speeds of metabolic change during their clinical timeline (**Figure 2g-h**), and sought to identify a clinical parameter that would be associated with that change (**Figure 2i**). We discovered that the overall speed of metabolic change along the whole timeline of the patient was related with the overall disease severity of the patients and whether the patient was hospitalized **(Figure 2j)**. This observation suggests that higher rates of metabolic changes over time are an index of the complex interactions between viral infection, treatment, and individual host response, and may translate into (or reflect) a worse overall outcomes for the patients. Taken together, our pseudo-temporal analysis of the metabolomics dataset revealed a dynamic landscape of metabolic change within patients over time, which can be used to measure disease progression in near-real time.

### Integration of immune and metabolic data for patient stratification

Since metabolic requirements underpin immune activation^47,48^ which is needed for response to infection^49–51^, and immune effectors are known to regulate key enzymes in lipid metabolism ^52,53^ we sought to uncover the relationship between metabolite abundance and immune system composition by performing regularized regression on the NMR metabolomics and flow cytometry immune profiling datasets^54^ (**Figure 3a** and **Figure S6**). This resulted in a map of interactions between metabolites and immune populations, of which Figure 3a illustrates the strongest. Interactions between immune and metabolic variables could be largely categorized in two groups: i) positive association (**Figure 3a** red in the heatmap): immune variables changing in the same direction as metabolic variables; ii) negative association (**Figure 3a** blue in the heatmap): increase in metabolic variables correlated with decrease in immune variables or vice-versa. For example, the decrease in Albumin levels during COVID-19 was matched with the increase in polymorphonuclear myeloid-derived suppressor cells (PMN-MDSC); total T-cell abundance was related to the fraction of medium size HDL, where both variables decrease with COVID-19 severity. The immune checkpoint inhibitors Lag3 and Tim3 which we previously described increasing with COVID-19 severity showed interactions with the fraction of cholesterol in LDL particles, themselves decreasing with COVID-19 severity. While many of these potential interactions are not yet described, and many are indirect, there are also specific examples of direct interactions, such as HDL interference with the potential of T cells to produce some cytokines, through a proposed mechanism of direct binding^55^. The relationships between the two datasets made us hypothesize that it could be possible to establish a patient-centric view of the immune-metabolic landscape during COVID-19. Towards that end, we employed regularized Canonical Correlation Analysis (rCCA) to integrate both NMR and flow cytometry datasets in a common latent space (**Figure 3b**). In this new space, samples clustered based on disease severity and its associated clinical outcomes regardless of dataset origin (**Figure 3b**). This allowed us to build a novel way to stratify patients based on both immune and metabolic data by hierarchical clustering of the pairwise similarity between patient samples (**Figure 3c**). In this classification we could identify six groups: one with predominantly healthy samples (11%); two groups of patients with mild disease (10 and 12% respectively); two groups mostly containing patients with severe disease (one with samples collected close to symptom onset, and the other later (11 and 24% respectively); and finally a group of samples from mostly convalescent patients (32%) (**Figure 3c**). The six groups were characterized by distinct clinical parameters and abundance of immuno-metabolic species (**Figure 3d**). For example, the two groups of mild disease could be distinguished by distinct BMI, liver enzyme levels, and triglyceride content of lipoproteins. Additionally, the “late” severe disease group had creatinine levels markedly higher than the “earlier” severe group, as opposed to B-cell expression of immunoglobulins G and M (IgG/IgM) which was highest in “early” disease and later decreased (**Figure 3d**).

**Figure 3:**
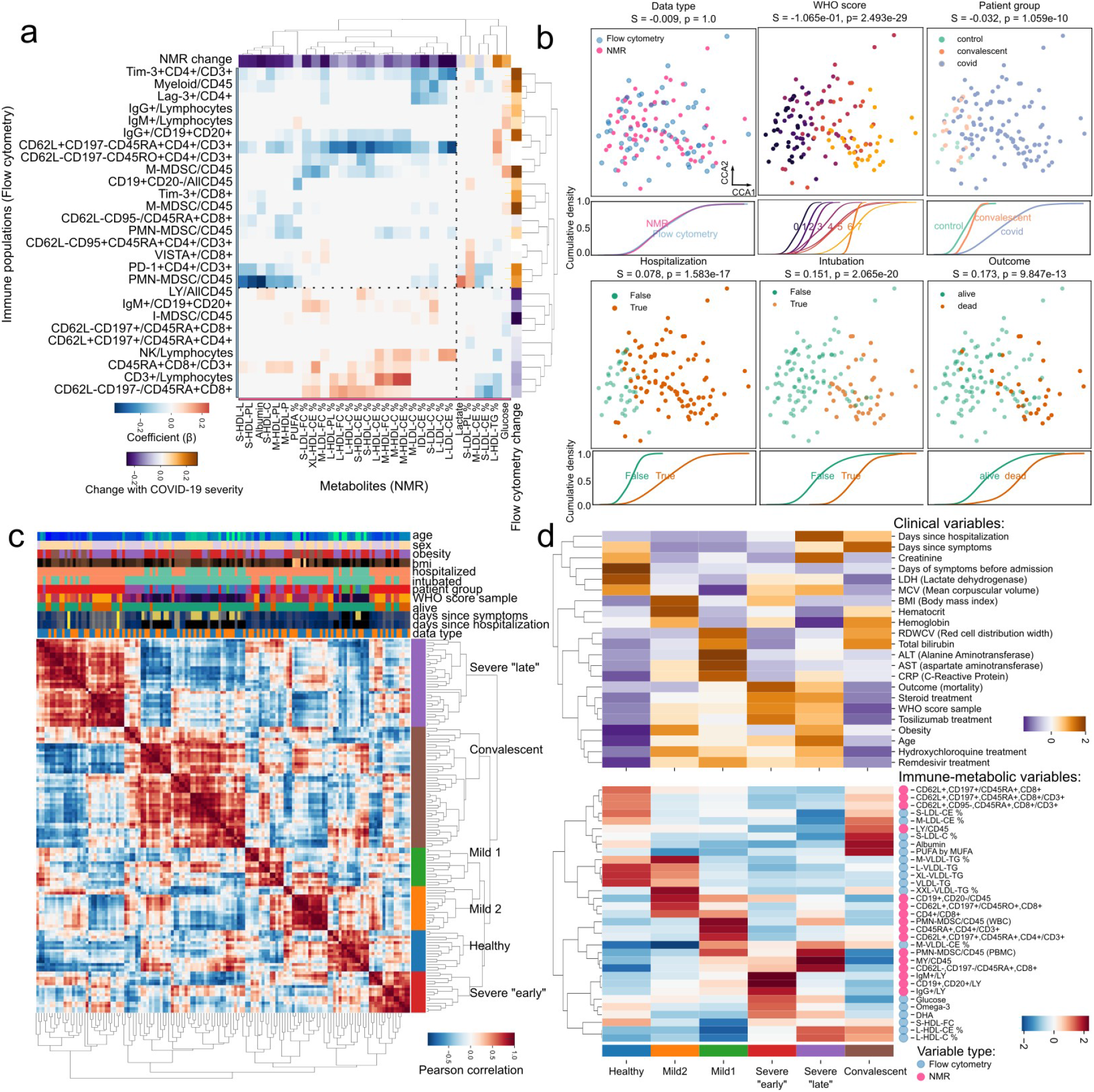
Joint immune-metabolic analysis of COVID-19 patients empowers novel patient stratification strategies. **a**) Heatmap of the relationship between metabolic (x-axis) and immune variables (y-axis). Only the 30 variables with most variance are shown per dataset. **b**) Integration of immune and metabolic data into a joint embedding. Each square panel demonstrates the distribution of samples dependent on clinical factors, and below the cumulative distribution function of each class along the first dimension. We provide silhouette scores (S) for how good the classes are separated and their significance through an ANOVA test (p). **c**) Pairwise correlation heatmap showing the similarity between samples based on immune-metabolic data. The hierarchical clustering dendrogram illustrates the newly discovered patient groups. Axis rows and columns are the same. Values of clinical parameters for every sample are illustrated above the heatmap. **d**) Association of sample groups with clinical (top) and immune-metabolic variables (bottom). Values have been row-wise Z-score transformed to account for the heterogeneous nature of the variables.

## Discussion

Here we present longitudinal immuno-metabolic data on a cohort of COVID-19 patients representative of the whole range of disease severity. We show that patient metabolism during disease is quite dynamic, reflecting disease progression and treatment. Consistent with a previous report, increased markers of systemic inflammation correlated with COVID-19 severity^25^. More importantly, we identified a deep alteration of the lipoprotein particles levels and composition: increased triglyceride content and VLDL, decrease of HDL, percentage of cholesterol/cholesteryl esters in HDL, and IDL were associated with severe disease. Previous studies have proposed a decrease in cholesterol and increase in triglycerides as markers of severe COVID-19^24,44^. In our study we confirm these findings and further describe the deep modification in the lipid metabolism and composition of lipoprotein and fatty acid associated with the disease. This is reminiscent of a metabolic state known to predispose to cardiovascular disease^56–58^ and could be related to the thrombotic events observed in COVID-19 patients with severe disease^2,4,5,59,60^. Furthermore, we develop tools to precisely monitor patient trajectories using metabolic data, enabling risk assessment on a continual fashion, and a novel patient stratification strategy. We must nonetheless acknowledge the following limitations to our study: i) our cohort is relatively small especially in comparison with large repositories such as the UK biobank; ii) our cohort is also skewed to have more patients with longitudinal follow-up for patients with severe disease - this is at least in part due to the natural dynamic severe disease having a longer recovery period; iii) our analysis of the interaction between the immune system and metabolome is purely correlational, as we can’t infer causality between the presence or activity of an immune cell type with the abundance of a metabolite.

It is plausible that cytokine modulation of key metabolic enzymes or energy usage by the immune system during acute infection are a major source of the metabolic changes associated with COVID-19 progression^61^. It has been shown for COVID-19 specifically that in T-cells cholesterol interacts with sphingolipids in membrane rafts in a manner that is dependent on the saturation state of the fatty acids^29^, and more generally that lipid raft formation has a crucial role in the cytotoxic activity of CD8 T-cells^62–64^. The increase in triglyceride composition of lipoprotein particles and their saturation state we observed with increased disease severity could result in altered immune function. Evidence for that has been seen in the regulation of immune checkpoint proteins such as CTLA4 in MDSCs by intracellular PUFA levels in cancer models^65^. Another example is the shift between energy sources in T effector cells from glucose to aminoacids which is required for proliferation and cytotoxic activity^49–51^.

Additional evidence of immune influence on metabolism in our data is the fact that tocilizumab - a neutralizing antibody of the pro-inflammatory IL-6 - partially rescues the effect of disease severity at the metabolic level (**Figure 1g**). This reinforces the idea that metabolic changes during COVID-19 are likely to be at least partially driven by the immune system either directly through regulation of key metabolic enzymes by cytokines, energy consumption of cytokine-secreting cells, or by the effect of immune cells on other tissues. At the same time, in our study BMI had a negligible influence on disease severity (**Figure S2a**), and that biomarkers for liver function such as AST and ALT did not show significant association with disease severity (**Figure S3b-c**), making nutrition, obesity and liver dysfunction unlikely candidates to explain metabolic change during COVID-19. Nonetheless, the contribution of these and other factors should be further explored in future studies with larger sample sizes.

The immune-metabolic crosstalk taking place during COVID-19 progression suggests the future potential use of metabolites to control disease through direct modulation of specific steps of lipid metabolism at the immune level. In that light, having precise methods for disease monitoring that capture both metabolism and immune system states would be extremely useful. In this study we develop a method for patient monitoring using NMR spectroscopy of metabolites from blood sera (**Figure 2**) that is quantitative, does not rely on thresholds, and can be interpreted in terms of patient risk at any given time during the patient’s clinical trajectory. Further development of our approach of joint immuno-metabolic classification of overall patient trajectories (**Figure 3**) could be used early in the course to tailor patient care and maximize allocation of medical resources.

Collectively, our study unveils the considerable metabolic disarray during COVID-19 progression which could open avenues for the development of metabolic-based therapies. Further, by leveraging immuno-metabolic high-dimensional data, we provide novel methods for precise disease monitoring and stratification in order to effectively tailor clinical care to COVID-19 patients.

## Methods

### Human studies

Blood serum samples were collected at the New York Presbyterian Hospital/Weill Cornell Medicine. Experiments using samples from human subjects were conducted in accordance with local regulations and with the approval of the IRB at the Weill Cornell Medicine. No statistical methods were used to pre-determine sample size.

### Targeted metabolomics with nuclear magnetic resonance

Analytes were quantified from plasma samples using targeted high-throughput NMR metabolomics (Nightingale Health Ltd.). 249 measures are produced, with 148 in absolute molar quantification. These include lipids, lipoprotein subclasses, fatty acids and their saturation, several low-molecular weight metabolites (amino acids, ketone bodies and glycolysis metabolites), as well as a set of clinically validated biomarkers associated with different metabolic pathways relevant to human physiology.

### Analysis of nuclear magnetic resonance data

In order to categorize the NMR analytes biophysically and functionally, we used data distributed by the *ggforestplot* package^34^ (https://github.com/NightingaleHealth/ggforestplot) and complemented them with variables representing lipoprotein particle size and density according to the variable names. Values of replicability per analyte were extracted from measurements of technical replicates performed by Nightingale Health Ltd. publicly available at: https://biobank.ndph.ox.ac.uk/showcase/showcase/docs/nmrm_app2.pdf. Summary statistics for metabolite species abundance at population scale were obtained from the publicly available resource showcase of the UK biobank^42^ (https://biobank.ndph.ox.ac.uk/ukb/) by querying field IDs 23400 to 23578. In order to build data-driven groups of variables, we used a standardized and centered matrix of features with absolute measurements only, and computed a nearest neighbor graph using 15 neighbors as the size of the local neighborhood (*scanpy*.*pp*.*neighbors*). These were used as input for UMAP (*scanpy*.*tl*.*umap*) and clustered using the Leiden algorithm (*scanpy*.*tl*.*leiden*), both with default parameters using Scanpy^66^.

To identify variables associated with COVID-19 severity, we performed linear regression using a mixed effect model. The model used age, gender, and body mass index (BMI) as covariates, with the WHO score per sample as the dependent variable, and fixed effects for each patient. To identify variables associated with tocilizumab treatment, we performed linear regression with a generalized linear model. The dependent variable was the time in days since treatment began, and only samples of patients which received treatment were included. Covariates of age, gender, and body mass index (BMI) were also used. We ensured there was no collinearity between predictors by measuring their variance inflation factor using statsmodels. We fit the models for all variables, inspected the distribution of residuals and for the mixed effect model also compared the estimated coefficients to a generalized linear model with no blocking on patient, and to models not incorporating the covariates. We found that the estimated effect of COVID-19 severity between these models was highly similar (r^2^ = 0.985). *p*-values were corrected for multiple testing using the Benjamini-Hochberg FDR method. To assess whether the group of features significantly associated with COVID-19 severity or tocilizumab treatment were enriched in any particular biophysical and functional classes, we performed enrichment analysis using parametric analysis of gene set enrichment^67^ (PAGE) as implemented in https://github.com/afrendeiro/page-enrichment.

### Generation of a latent space for precision disease monitoring

To establish a latent space embedding using the metabolomics data, we performed spectral embedding of the metabolomics data (sklearn.manifold.SpectralEmbedding). We also compared the results of this method to the following methods for dimensionality reduction: principal component analysis (PCA), non-negative matrix factorization (NMF), multidimensional scaling (MDS), non-linear dimensionality reduction through isometric mapping (Isomap), t-distributed stochastic neighbor embedding^68^ (t-SNE), uniform manifold approximation and projection (UMAP), as implemented in scikit-learn, diffusion maps (DiffMap) as implemented in Scanpy^66^, and minimum-distortion embedding^66,69^ (MDE) from the *PyMDE* package. Spectral embedding produces exactly the same results as diffusion maps (DiffMap) with default parameters if the input matrix is standardized and centered. To order variables along a gradient within the derived latent space across its two dimensions, we correlated the original features with each latent vector, scaled each to the unit range and multiplied the values of dimension 1 and 2. Then, to order samples along this gradient, we simply computed the correlation of each sample with the previously derived vector.

Inference of clinical parameters distribution within the latent space was done as previously^70^ : two bivariate gaussian kernel density estimators were fitted on the coordinates of the samples with the difference being that one was weighted by the respective value of the sample in the clinical parameter. The final values are given by the difference between the two estimators. The compound measure of relative risk is the average of these estimations for the WHO score, hospitalization, intubation, and death.

To generate a vector field of patient movement through the latent space, we extracted vectors representing the movement of each sample at each timepoint by dividing the euclidean distance between points by the time between each two consecutive timepoints. Then, we interpolated these values across the two-dimensional latent space (*scipy*.*interpolate*.*griddata*). The total velocity of each patient in the space was calculated as the total distance over the length of the timeline (first to last NMR sample). To derive a score of COVID-19 severity for each sample, we also separated features dependent on the sign of the coefficients of the mixed effects model and calculated the difference in the mean of up-regulated features and mean down-regulated features scaled by their relative size.

### Joint analysis of nuclear magnetic resonance and flow cytometry datasets

In order to understand the relationship between metabolic variables and immune populations, we performed Ridge regression between the NMR and flow cytometry datasets with hyperparameter optimization using random search cross-validation (*sklearn*.*model_selection*.*RandomizedSearchCV*) for the alpha parameter sampled from a log-uniform distribution with parameters a = 1e-20, and b = 1 for 1000 iterations. The coefficients of the best model were highly regularized (alpha = 0.979196) and were used to represent the relationship between metabolic and immune population variables.

To produce a joint embedding of metabolic and immune data for each patient timepoint, we employed regularized canonical correlation analysis^71^ (RCCA) in the Python implementation *pyrcca*. We performed hyperparameter optimization with grid search cross validation using a number of canonical components between 4 and 8, and a regularization parameter between 1e-3 and 1e3. The best number of canonical components was 6 and the regularization parameter was 90. The separation of groups of samples dependent on clinical parameters was assessed with a silhouette score and an analysis of variance (ANOVA) test on the first 2 canonical components only.

To produce a stratification of patients based on the joint projection of the two datasets in the RCCA space by correlating samples in a pairwise fashion and extracting the first 6 splits of a dendrogram derived from hierarchical clustering of the correlation coefficients. The association of clinical or immune-metabolic variables with the derived patient groups was performed by fitting a linear model explaining those variables using the patient groups. In the case of immune-metabolic data, only the top 3 variables per group were chosen for visualization.

#### Software used

Python version 3.8.2, numpy^72^ 1.21.0, scipy^73^ 1.7.0, statsmodels^74^ 0.12.2, scikit-learn^75^ 0.24.2, scanpy^66^ 1.8.0, pymde^69^ 0.1.12, *pingouin*^*76*^ 0.3.12, and pyrcca^71^ 0.1.

## Data Availability

Clinical and demographic annotation of the samples is provided as Table S1. The full NMR metabolomics dataset is provided as Table S2 and Table S3.Previously publishedflow cytometry data54are available at the following URL: https://github.com/ElementoLab/covid-flowcyto

## Data availability

Clinical and demographic annotation of the samples is provided as Table S1. The full NMR metabolomics dataset is provided as Table S2 and Table S3. Previously published flow cytometry data^54^ are available at the following URL: https://github.com/ElementoLab/covid-flowcyto

## Code availability

Source code for the full data analysis of the study is available at the following URL: https://github.com/ElementoLab/covid-metabolomics

## Acknowledgements

A. F. R. is supported by a NCI T32CA203702 grant. O.E. is supported by NIH grants UL1TR002384, R01CA194547, and Leukemia and Lymphoma Society SCOR 7012-16, SCOR 7021-20 and SCOR 180078-02 grants. C.K.V. was supported by NIAID K08 AI132739 and a Potts Memorial Foundation Award. We would like to thank Karsten Suhre for his expertise on metabolic analysis.

## Author contributions

G.I., O.E., and M.S. planned the study; C.K.V., H.S., S.K. provided samples and clinical data; L.V.C, M.T.C. processed samples; A.F.R. performed analysis of the data with contributions from J.K.; G.I., O.E., and M.S. supervised the research. A.F.R., L.V.C., J.K., O.E., and M.S. wrote the manuscript with contributions from all authors.

## Supplementary figures

**Figure S1:**
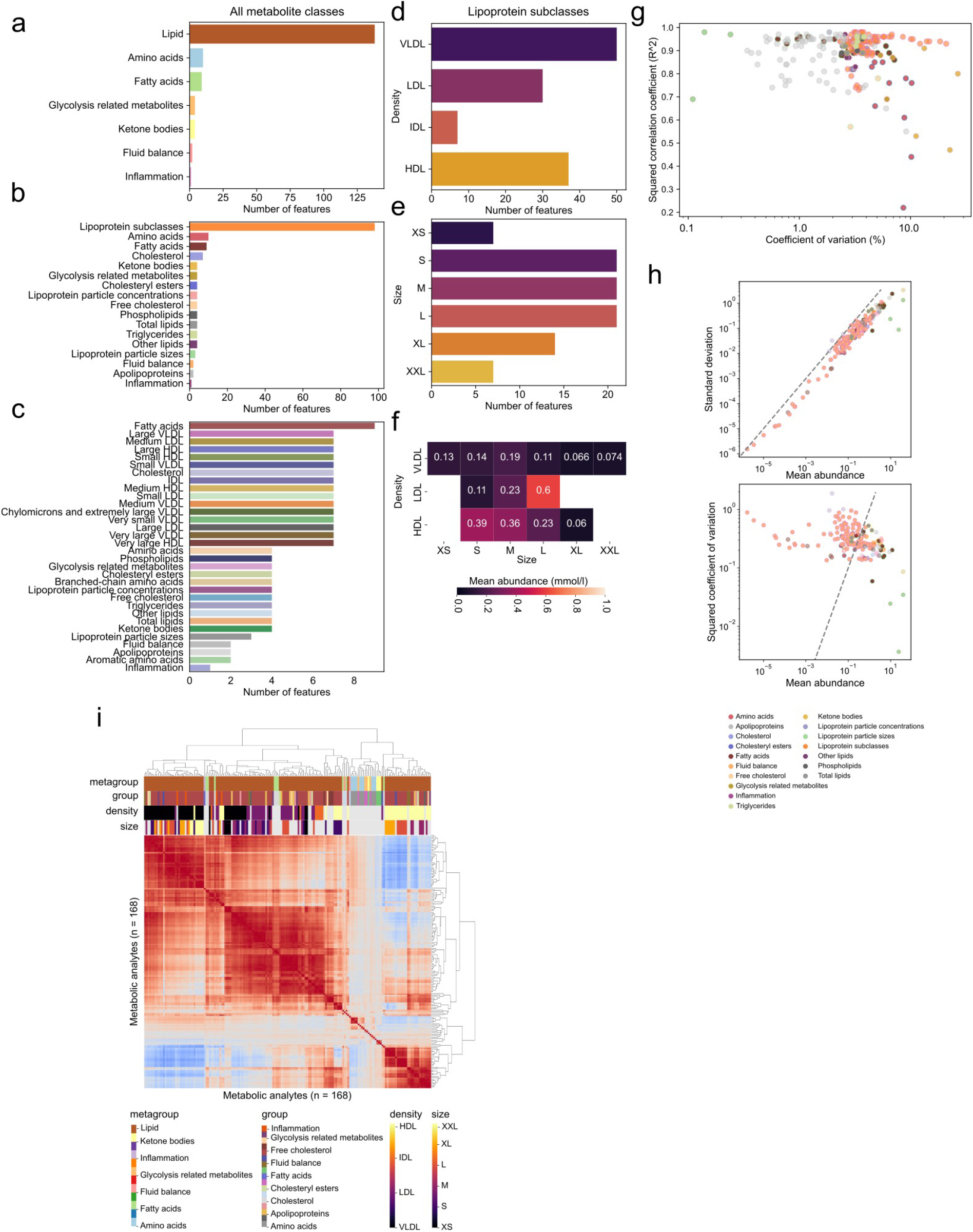
Characterization of the NMR metabolomics panel. **a-c**) Composition of the panel dependent on the biophysical characteristics of the analytes from a) to c) with increased granularity. **d-e**) Composition of the lipoprotein particle variables in the panel depending on their density d) and size e). **f**) Absolute abundance of metabolites depending on their density or size. **g**) Measures of reproducibility and signal-to-noise for all metabolites in the panel. **h**) Relationship between mean and variance for all variables in the panel. **i**) Pair-wise correlation of metabolite abundance.

**Figure S2:**
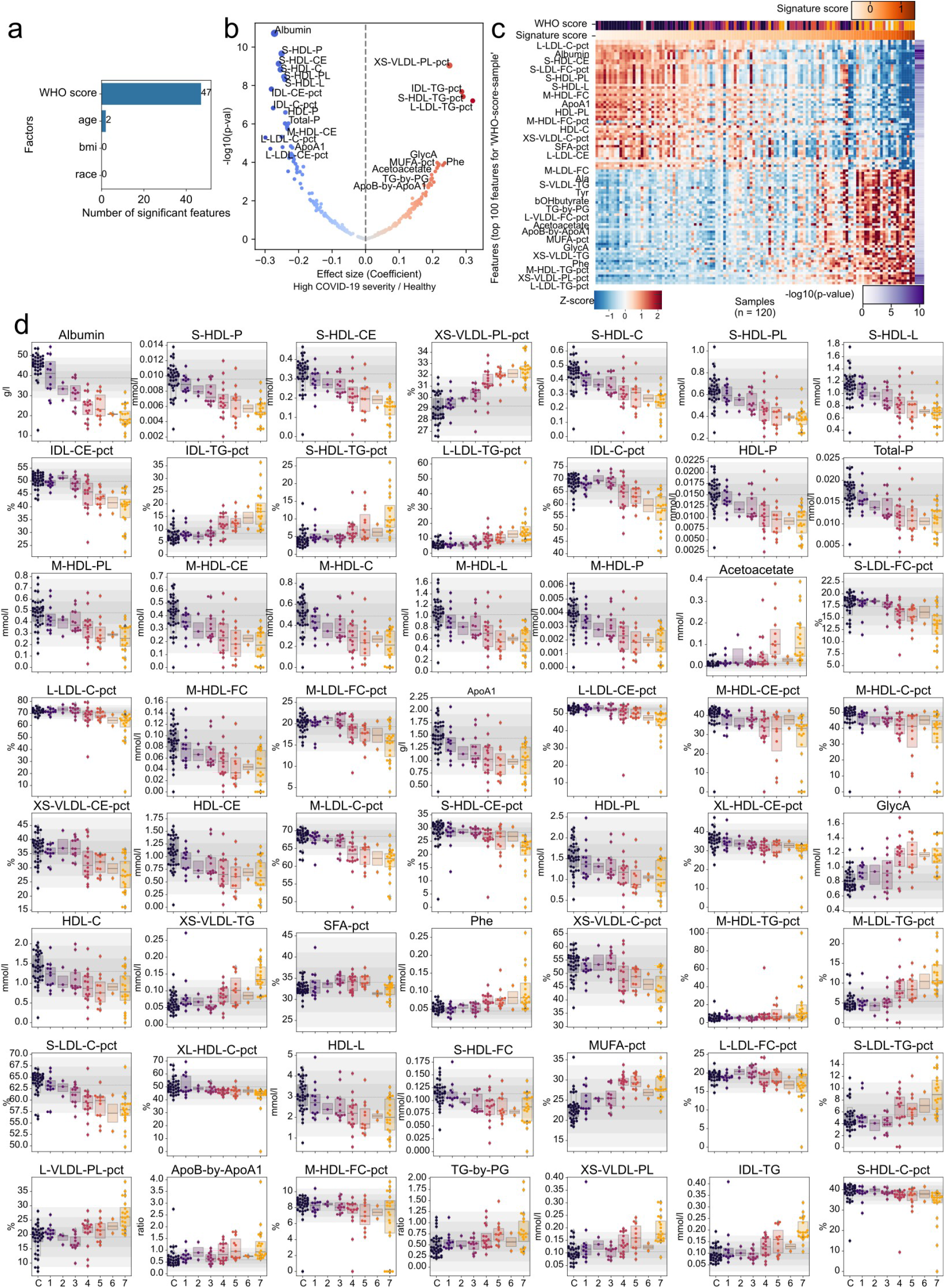
Metabolic changes associated with COVID-19 severity. **a**) Number of significant (p < 0.05 FDR) variables for a joint model of COVID-19 severity, patient age, BMI, and race. **b**) Volcano plot of changes in metabolites associated with COVID-19 severity. **c**) Heatmap of relative metabolite abundance for all samples where the axes have been sorted by the amount of change. **d**) Abundance of metabolites with significant association with COVID-19 severity depending on WHO score.

**Figure S3:**
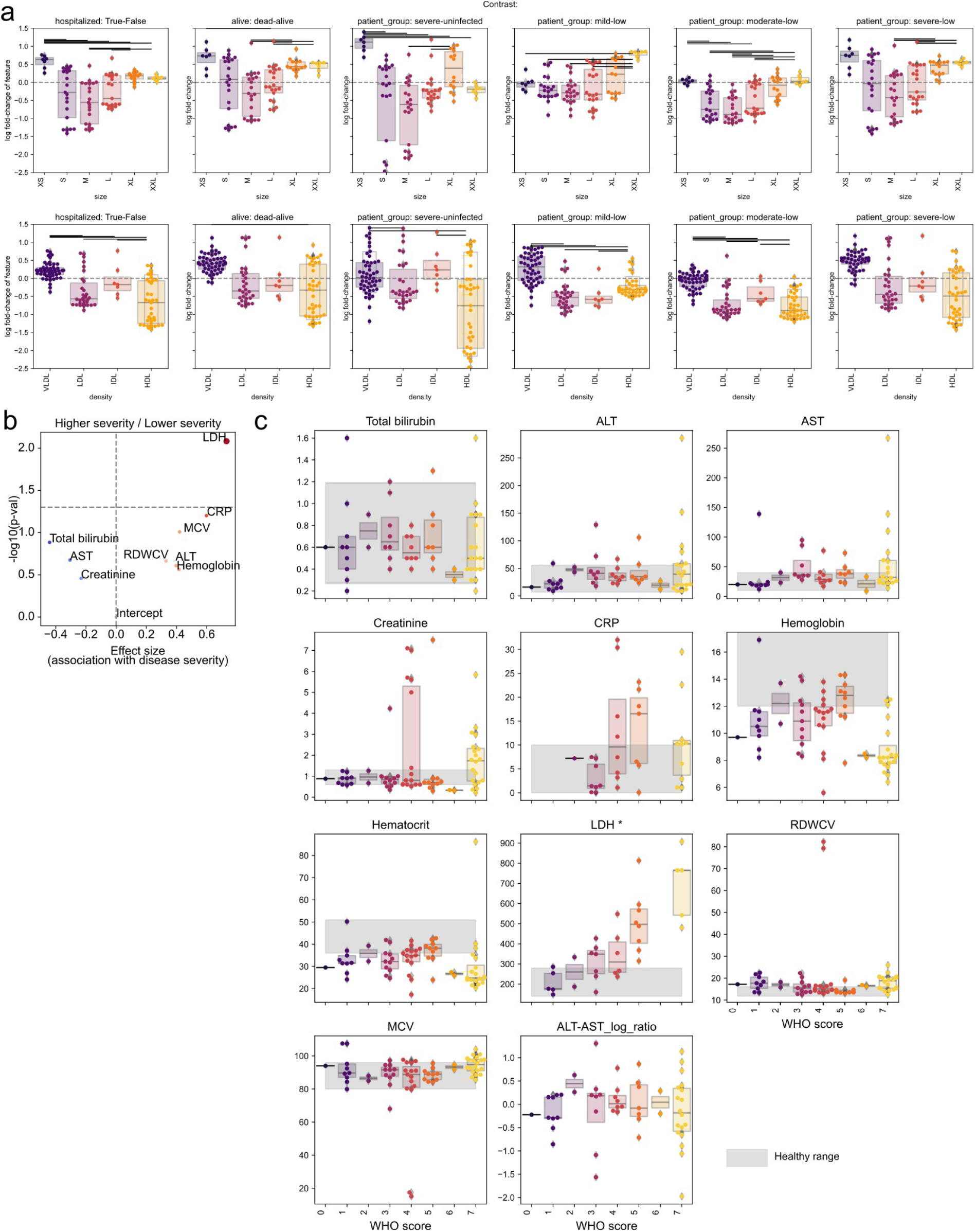
Metabolic and clinical association of disease severity. **a**) Distribution of log fold-changes in lipoprotein particle metabolites depending on their size (upper row) or density (lower row). The coefficients represent the change associated with hospitalization, death and disease severity. **b**) Volcano plot of clinical variables associated with COVID-19 severity in our cohort. **c**) Distribution of clinical parameters in the samples dependent of COVID-19 severity. The horizontal grey areas represent a healthy range for each parameter.

**Figure S4:**
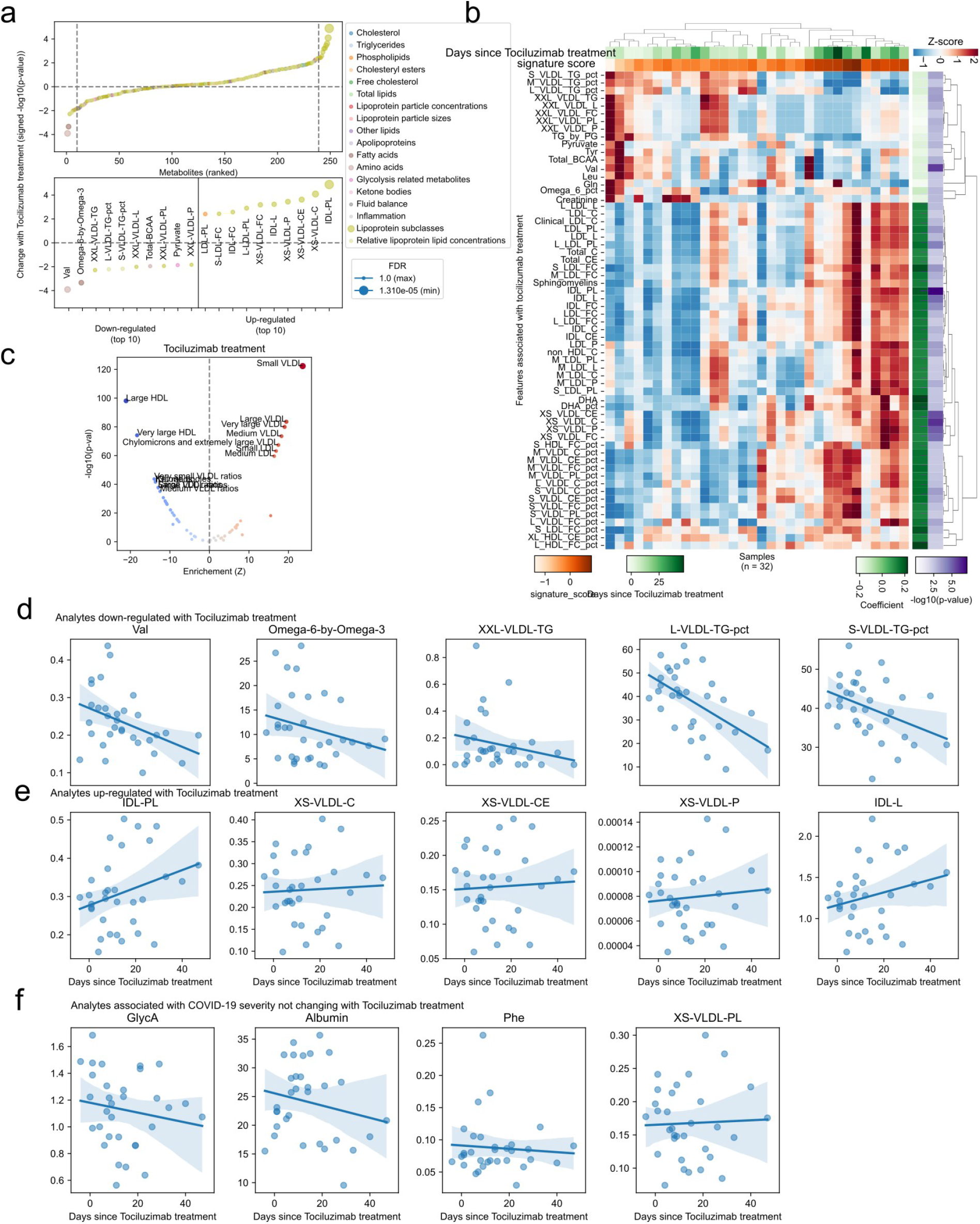
Metabolic changes associated with tocilizumab treatment in COVID-19. **a**) Association of metabolite abundance with the time of tocilizumab treatment for all metabolic species (upper panel). The lower panel illustrates the 10 metabolites most associated in each direction. **b**) Heatmap of metabolites significantly associated with tocilizumab treatment for samples of patients that have been treated. Volcano plot of clinical variables associated with COVID-19 severity in our cohort. **c**) Enrichment of metabolite classes in the change with tocilizumab treatment. **d-f**) Abundance of metabolites with discordant d), concordant e) or indifferent d) change between COVID-19 severity and tocilizumab treatment for treated patients.

**Figure S5:**
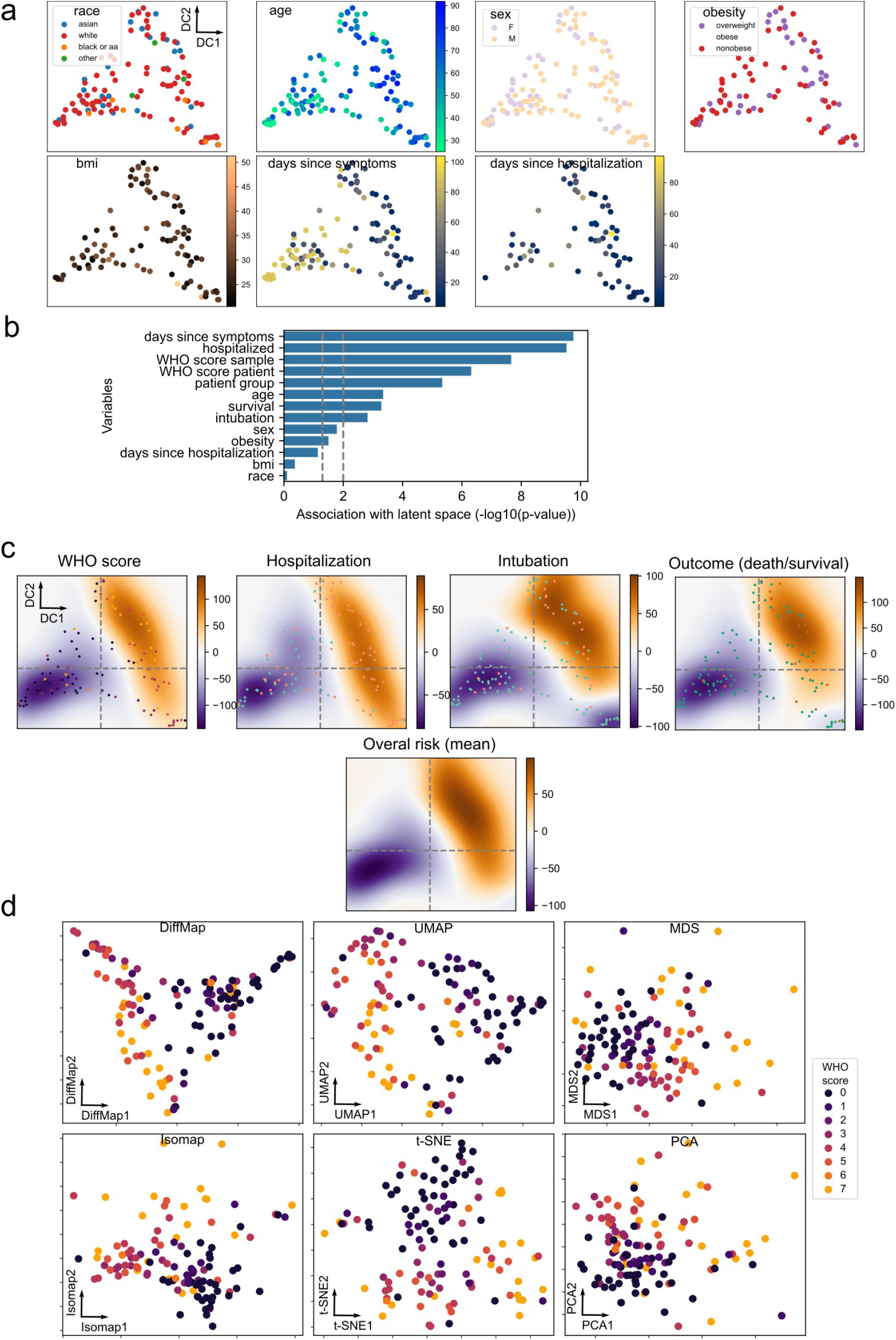
Metabolic and clinical association of disease severity. **a**) Latent space as in Figure 2a, but illustrating the distribution of additional clinical factors. **b**) Association analysis of clinical variables with the latent space axes. *p*-values have been adjusted with the Benjamini-Hochberg FDR method. **c**) Difference between bivariate kernel density estimates that have been weighted with the clinical parameters of the samples. The overall risk is the mean of the four clinical parameters in the first row of plots. **d-f**) Latent space embeddings of metabolomic data using alternative methods. Samples have been colored by the WHO score scale.

**Figure S6:**
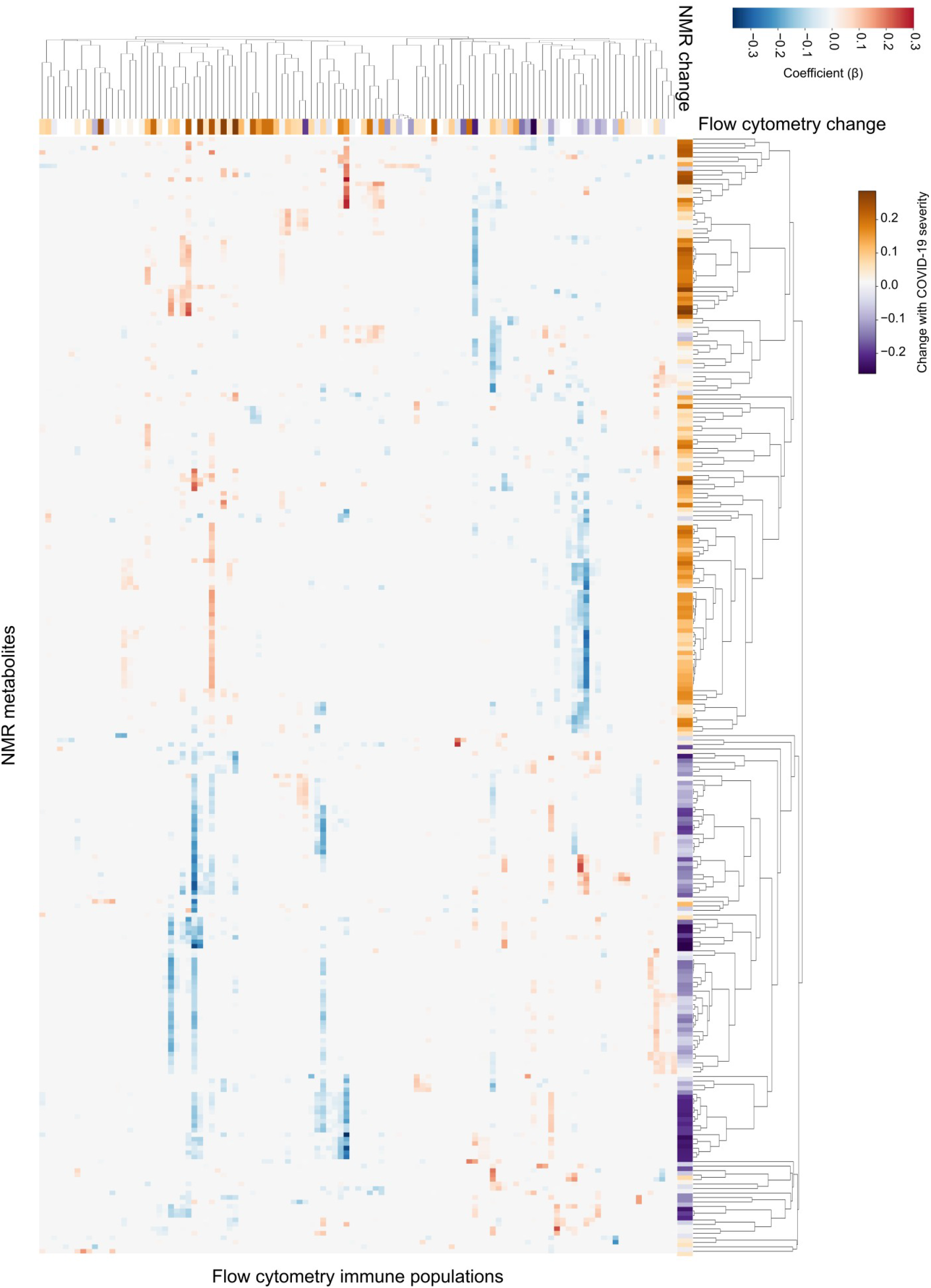
Interactions between immune and metabolic variables during COVID-19. Heatmap of the relationship between all immune (x-axis) and metabolic variables (y-axis). The change of each feature in relation to COVID-19 severity is displayed in a purple-to-gold heatmap.

